# The EEG multiverse of schizophrenia

**DOI:** 10.1101/2020.12.21.20248665

**Authors:** Dario Gordillo, Janir Ramos da Cruz, Eka Chkonia, Wei-Hsiang Lin, Ophélie Favrod, Andreas Brand, Patrícia Figueiredo, Maya Roinishvili, Michael H. Herzog

**Author notes:** ***Corresponding author:*** Dario Gordillo, Laboratory of Psychophysics, Brain Mind Institute, School of Life Sciences, École Polytechnique Fédérale de Lausanne (EPFL), CH-1015 Lausanne, Switzerland. These authors contributed equally (Dario Gordillo, Janir Ramos da Cruz).

## Abstract

Research on schizophrenia typically focuses on one paradigm, for which clear-cut differences between patients and controls are established. Great care is taken to understand the underlying genetical, neurophysiological, and cognitive mechanism, which eventually may explain the clinical outcome. One tacit assumption of these *deep rooting* approaches is that paradigms tap into common and representative aspects of the disorder. Here, we analyzed the resting-state electroencephalogram (EEG) of 121 schizophrenia patients and 75 controls. Using multiple signal processing methods, we extracted 194 EEG features. Sixty-nine out of the 194 EEG features showed a significant difference between patients and controls indicating that these features detect an important aspect of schizophrenia. Surprisingly, the correlations between these features were very low, suggesting that each feature picks up a different aspect of the disorder. We propose that complementing *deep* with *shallow* rooting approaches, where many roughly independent features are extracted from one paradigm (or several paradigms), will strongly improve diagnosis and potential treatment of schizophrenia.

## 1. Introduction

Schizophrenia patients show strong abnormalities in many domains including personality, cognition, perception, and even immunology. In many experimental paradigms, the differences between patients and controls have large effect sizes, indicating that important aspects of the disease are detected. This provokes two questions: what do these abnormalities have in common and how representative are they for the disease? For example, patients exhibit strong deficits in cognition, such as in working memory tasks (Meyer-Lindenberg et al., 2001), which are attributed to abnormalities of cortico-cerebellar-thalamic-cortical circuits (Andreasen et al., 1998). Patients show also diminished skin flushing with the niacin skin test (Rybakowski and Weterle, 1991), which is attributed to dysfunctional phospholipase A2 arachidonic acid signaling (Messamore, 2012). How do the working memory deficits correspond to deficits in skin functioning? Very few studies have correlated deficits with each other (Braff et al., 2007a, 2007b; Dickinson et al., 2011; Price et al., 2006; Seidman et al., 2015; Toomey et al., 1998). The Consortium on the Genetics of Schizophrenia studied neurocognitive and neurophysiological abnormalities in schizophrenia patients with a battery of 15 paradigms (Seidman et al., 2015). They found that neurocognitive measures shared a significant amount of variance while neurophysiological measures were almost entirely independent. Price and colleagues (2006) studied four candidate electrophysiological endophenotypes of schizophrenia (mismatch negativity, P50, P300, and antisaccades). Even though patients *and* their family members showed deficits in each of these endophenotypes, the features were largely uncorrelated.

Here, we took another road. Instead of comparing different paradigms, we analyzed the very same data of the very same patients with different electroencephalogram (EEG) analysis methods, including many that have shown strong atypical patterns in patients (Andreou et al., 2015; Boutros et al., 2008; da Cruz et al., 2020; Di Lorenzo et al., 2015; Kim et al., 2000, p. 200; Nikulin et al., 2012; Sun et al., 2014; Uhlhaas and Singer, 2010). Resting-state EEG features revealing significant differences between schizophrenia patients and controls are thought to reflect brain mechanisms linked to important aspects of the disorder. For example, schizophrenia patients exhibit reduced long-range temporal correlations (LRTC) in the alpha and beta frequency bands (Nikulin et al., 2012). These responses were suggested to reflect excessive switching of neuronal states in patients. Patients also have shown atypical patterns in the dynamics of EEG microstates classes C and D (da Cruz et al., 2020; Rieger et al., 2016). Microstates abnormalities have been proposed to correspond to imbalances in attentional and information processing in schizophrenia. Schizophrenia patients have shown increased power in the delta, theta, and beta frequency bands (Venables et al., 2009). Increased beta power was suggested to reflect cortical hyperexcitability, increased power in the delta and theta bands were proposed to relate to atypical dopaminergic function, to name a few examples. All these results, individually, suggest that each EEG feature captures important aspects of schizophrenia. But how representative are these abnormalities of the disorder? Does a patient showing abnormal microstate dynamics also show deficits in LRTC, or in other EEG features? Even though all these atypical patterns in patients are obtained from the same EEG data, no study has evaluated how these EEG features relate to each other. This is not only the case for resting-state EEG studies, but constitutes the conventional approach in schizophrenia research. This approach is centered on drawing general conclusions about schizophrenia based on one paradigm, assumed to unveil common and representative aspects of the disorder.

Aiming to shed light on this EEG *multiverse* of schizophrenia, in this work, we analyzed the resting-state EEG data of 121 schizophrenia patients and 75 healthy controls with multiple methods. This allowed us to extract 194 EEG features, such as time-domain features, frequency-domain and connectivity features both in electrode and source space, and nonlinear dynamical features. Then, we correlated the features revealing group differences to evaluate how these abnormalities/deficits relate to each other. We also examined whether EEG features show adequate predictive power to clinical scales measuring key symptoms of schizophrenia. We propose that future studies in schizophrenia research should consider multiple features extracted from the same and/or different paradigms in order to improve diagnosis and potential treatment.

## 2. Material and Methods

### 2.1. Participants

Two groups of participants joined the experiment: schizophrenia patients (*n* = 121) and healthy controls (*n* = 75). All participants took part in a battery of tests comprising perceptual and cognitive tasks as well as EEG recordings. Data of 101 patients and 75 controls have already been published in different contexts (da Cruz et al., 2020a, 2020b; Favrod et al., 2018; Garobbio et al., 2021). Patients were recruited from the Tbilisi Mental Health Hospital or the psycho-social rehabilitation center. Patients were invited to participate in the study when they had recovered sufficiently from an acute psychotic episode. Thirty-five were inpatients and 86 were outpatients. Patients were diagnosed using the Diagnostic and Statistical Manual of Mental Disorders Fourth Edition (DSM-IV) by means of an interview based on the Structured Clinical Interview for DSM-IV, Clinical Version, information from staff, and study of patients’ records. Psychopathology of patients was assessed by an experienced psychiatrist using the Scale for the Assessment of Negative Symptoms (SANS) and the Scale for the Assessment of Positive Symptoms (SAPS). Out of the 121 patients, 106 were receiving neuroleptic medication. Chlorpromazine (CPZ) equivalents are indicated in **Table 1**. Controls were recruited from the general population in Tbilisi, aiming to match patients’ demographics as closely as possible. All controls were free from psychiatric axis I disorders and had no family history of psychosis. General exclusion criteria were alcohol or drug abuse, severe neurological incidents or diagnoses, developmental disorders (autism spectrum disorder or intellectual disability), or other somatic mind-altering illnesses, assessed through interview by certified psychiatrists. All participants were no older than 55 years. Group characteristics are presented in **Table 1**. All participants signed informed consent and were informed that they could quit the experiment at any time. All procedures complied with the Declaration of Helsinki (except for pre-registration) and were approved by the Ethical Committee of the Institute of Postgraduate Medical Education and Continuous Professional Development (Georgia). Protocol number: 09/07. Title: “Genetic polymorphisms and early information processing in schizophrenia”.

**Table 1.** Group average statistics (±SD)

|  | Patients | Controls | Statistics |
| --- | --- | --- | --- |
| Gender (F/M) | 22/99 | 39/36 | $\chi^2(1) = 24.702, p = 6.690e-7^b$ |
| Age (years) | $35.8 \pm 9.2$ | $35.1 \pm 7.7$ | $t(194) = 0.519, p = 0.604^c$ |
| Education (years) | $13.3 \pm 2.6$ | $15.1 \pm 2.9$ | $t(194) = -4.418, p = 1.657e-5^c$ |
| Handedness (L/R) | 6/115 | 4/71 | $\chi^2(1) = 0.013, p = 0.908^b$ |
| Illness duration (years) | $10.8 \pm 8.7$ | | |
| SANS | $10.1 \pm 5.2$ | | |
| SAPS | 8.6 ± 3.2 |  |  |
| CPZ equivalent<sup>a</sup> | 561.1 ± 389.4 |  |  |
| SANS - Scale for the Assessment of Negative Symptoms, SAPS - Scale for Assessment of Positive,<br>CPZ - chlorpromazine<br><sup>a</sup>Average CPZ equivalents calculated over the 106 Patients receiving neuroleptic medication<br><sup>b</sup>Pearson's chi-squared test<br><sup>c</sup>Two-sided independent samples *t*-test |  |  |  |

### 2.2. EEG recording and data processing

Participants were sitting in a dim lit room. They were instructed to keep their eyes closed and to relax for 5 minutes. Resting-state EEG was recorded using a BioSemi Active Two Mk2 system (Biosemi B.V., The Netherlands) with 64 Ag-AgCl sintered active electrodes, referenced to the common mode sense electrode. The recording sampling rate was 2048 Hz. Offline data were downsampled to 256 Hz and preprocessed using an automatic pipeline (da Cruz et al., 2018). Preprocessed EEG data were analyzed using multiple signal processing methods in the electrode and source space. In total, 194 EEG features were extracted (See Supplementary Table 1). Out of the 194 EEG features, 50 were obtained in the source space and 144 in the electrode space. For source space analysis, we defined 80 brain regions (40 per hemisphere) according to the AAL atlas (See Supplementary Table 4). See Supplementary Methods for a detailed description of the analysis methods.

### 2.3. Group comparisons

We compared patients’ and controls’ scores for each of the 194 EEG features. For each of the *J* variables (i.e., 64 electrodes, 80 brain regions, or 12 microstate parameters, depending on the number of variables of each EEG feature) of a given feature, we performed a two-way ANCOVA, with Group (patients and controls) and Gender (male and female) as factors and Education as a covariate. *P*-values for the effect of Group were corrected for *J* comparisons using False Discovery Rate (FDR; with an error rate of 5%). Group effects’ *η*^2^ were converted to Cohen’s *d*.

### 2.4. Pearson and partial least squares correlations

First, for each EEG feature that contained at least one variable showing a significant difference between patients and controls (after correcting for multiple comparisons), we selected the variable (i.e., electrode, brain region, or microstate parameter) with the biggest effect size to be the representative variable for that feature. Then, for patients and controls separately, we computed pairwise Pearson correlations between the representative variables of each significant EEG feature. Second, to quantify the overall relationship, i.e., the amount of *shared information*, between pairs of multivariate EEG features, we used Partial Least Squares Correlation (PLSC). PLSC generalizes correlations between two variables to two matrices (McIntosh et al., 1996; Tucker, 1958). The *shared information* can be quantified as the *inertia* common to the two features (Krishnan et al., 2011). The statistical significance of the inertia was assessed using a permutation test (Abdi and Williams, 2013; McIntosh et al., 2004). The inertia values were normalized. Hence, the normalized inertias (ℑ_*relative*_) ranged from 0 (the two EEG features are completely unrelated) to 1 (the two EEG features contain the same information). PLSC analysis was done for patients and controls separately. See Supplementary Methods for details.

### 2.5. Regression analysis

To evaluate whether EEG features predict psychopathology scores (SAPS and SANS) adequately, we used elastic net regression models (Zou and Hastie, 2005). Elastic nets can handle regression problems where the number of predictors is relatively large compared to the number of samples as well as multicollinearity (i.e., the predictors are not linearly independent) by combining the L1 and L2 penalties to achieve regularization. For each of the 194 EEG features (with all its variables), we built two regression models, one to predict SAPS scores and one to predict SANS scores. We performed 20 repetitions of a 3-fold nested cross-validation procedure. First, one third of the data (1 fold) was left out for validation (test set), while the remaining data (2 folds; train set) were used to find the optimal parameters, namely the amount of penalization and the compromise between L1 and L2 penalties, using 3-fold cross-validation. The model with the parameters leading to best performance in the train set was tested on the left-out data (test set). The entire procedure was repeated 20 times, with different allocations of the patients in the train and test sets. Using the same cross-validation procedure, i.e., 20 repetitions of a 3-fold cross-validation, we also evaluated predictive performance using a nonlinear random forest regression model, setting the maximum depth of the tree to 10 and the number of trees to 100. Random forests are meta estimators that average several decision trees trained on subsets of the dataset to improve accuracy and avoid overfitting. Prediction performance was calculated using the coefficient of determination (*R2*) and the root mean squared error (*RMSE*). The distribution of the prediction performance values was obtained from the 60 aggregated RMSE and *R2*, across repetitions of the procedure.

## 3. Results

### 3.1. Multiple EEG features reveal significant group effects

For 121 patients (22 females, 35.8 ± 9.2 years old, 13.3 ± 2.6 years of education) and 75 age-matched healthy controls (39 females, 35.1 ± 7.7 years old, 15.1 ± 2.9 years of education; Table 1), we extracted in total 194 features from the resting-state EEG recordings, including time-domain, frequency-domain, connectivity, and nonlinear dynamical features (Supplementary Table 1). Among the 194 EEG features, 69 (35.57%) showed significant differences between patients and controls with medium to large effect sizes (Cohen’s *d* varied from 0.463 to 1.037, **Figure 1**). Patients showed significantly reduced values in 24 out of the 69 EEG features revealing significant group differences (illustrated as negative effect size in **Figure 1**). Patients exhibited significantly higher values than controls in 45 EEG features.

**Figure 1.**
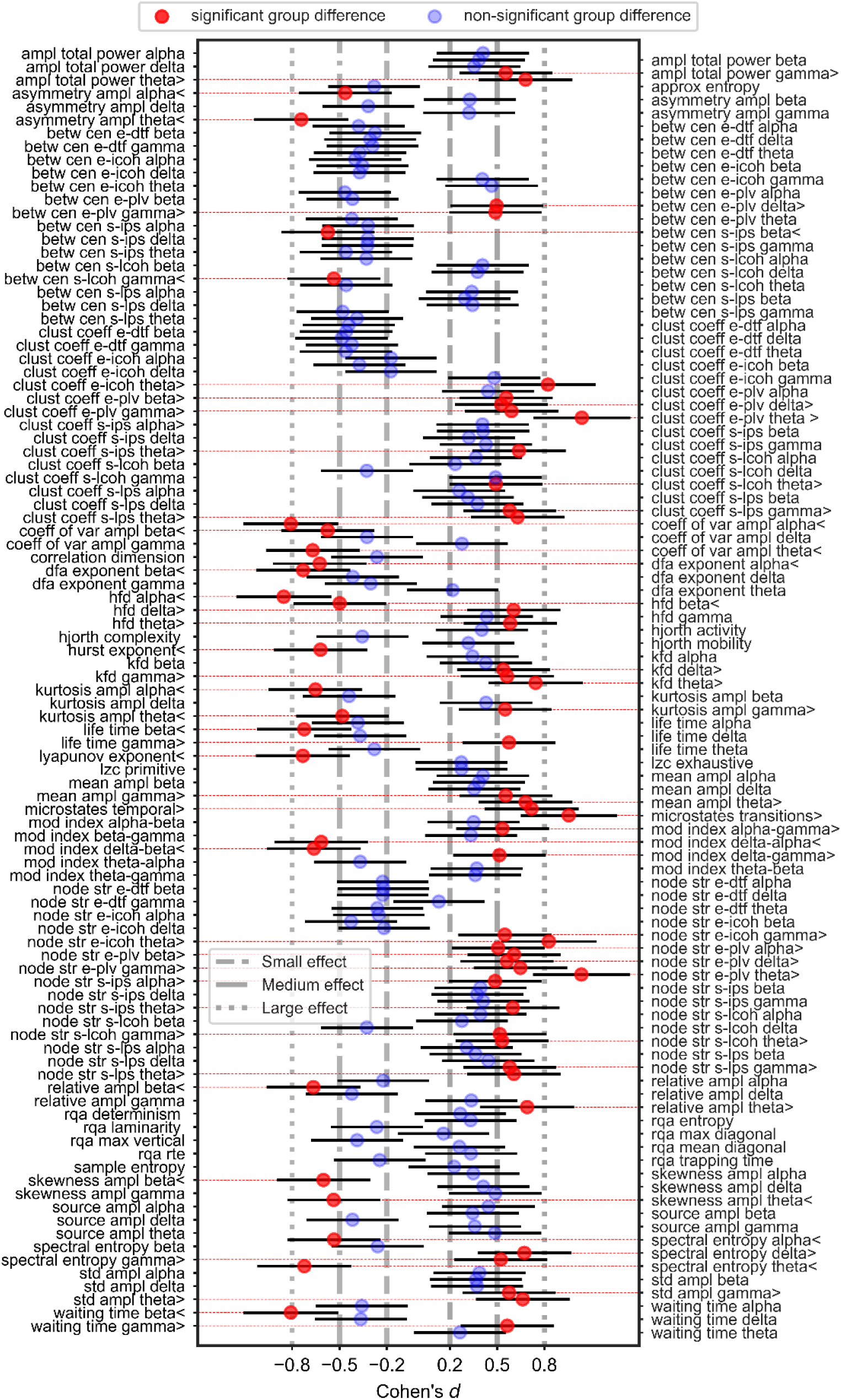
Effect size (Cohen’s *d*) of the group differences between patients and controls for each of the 194 EEG features. We took the values of the electrode, brain region, or microstate parameter with the largest effect size according to Cohen’s *d* (*η*^2^ values were converted to Cohen’s *d*) to be the representative variable for each feature. Significant group differences, after correction for multiple comparisons (using FDR), are depicted in red, with dotted red vertical lines serving as a guide to their labels. > and < were added to the feature labels to indicate if patients had significantly higher or lower values than controls, respectively. The non-significant effects are shown in blue. Error bars represent 95% confidence intervals (C.I.). A list with the abbreviations and the corresponding name of each feature is presented in Supplementary Table 1.

### 3.2. Correlations between EEG features

To evaluate to what extent features that showed significant group differences are sensitive to the same aspects of the disorder we computed Pearson’s correlations between pairs of features (**Figure 2**). As the representative variable for each feature, we took the values of the electrode, brain region, or microstate parameter that showed the largest group difference according to Cohen’s *d* (**Figure 1**). Surprisingly, we found that in the patients group only 36.49% of the pairwise correlations were significant at a level of 0.05 (without correcting for multiple comparisons). For the control group, only 26.73% of the correlations were significant. Since significance depends on the sample size, here, we focus on the magnitude of the correlation coefficients (*r*-values). In general, the magnitudes of the *r*-values were very low in both patients (0.055, 0.122, 0.251, for the 25^th^, 50^th^, and 75^th^ percentiles, respectively) and controls (0.059, 0.129, 0.242, for the 25^th^, 50^th^, and 75^th^ percentiles, respectively; **Figure 2**). Strong correlations were found mainly for pairs of very closely related features, such as between waiting-time statistics of gamma bursts (*waiting time gamma*) and life-time statistics of gamma bursts (*life time gamma*; *r*=0.836 and *r*=0.926, in patients and controls, respectively).

**Figure 2.**
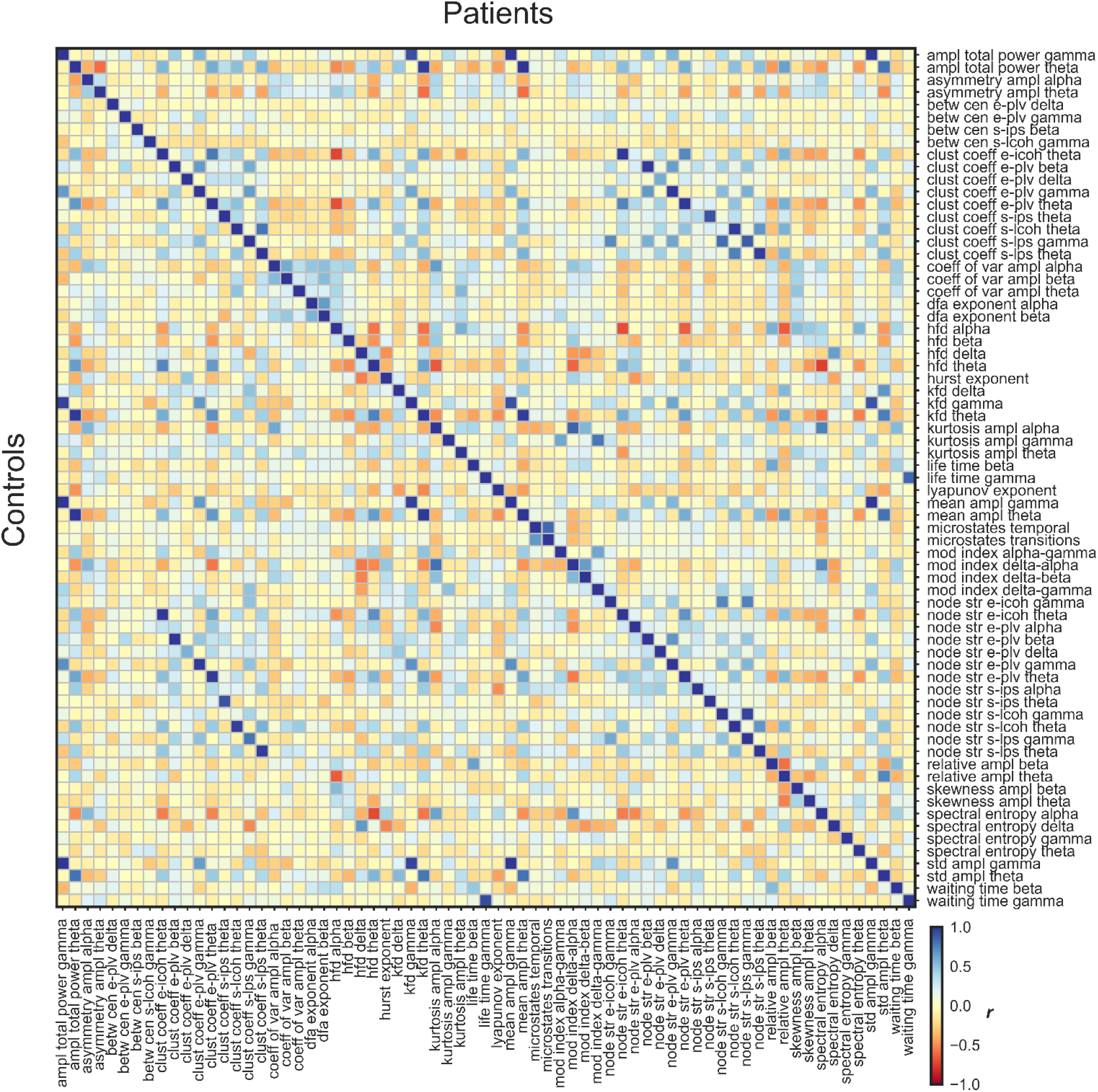
Pairwise correlations between the 69 EEG features that showed significant group differences between patients and controls. Patients’ *r*-values are presented in the upper triangle and controls’ *r*-values are shown in the lower triangle. Strong negative and positive *r*-values are depicted in red and blue, respectively, and *r*-values around 0 in yellow. For each feature, we used the values of the electrode, brain region, or microstate parameter that showed the largest effect size as the representative variable for the correlations. A list with the abbreviations and corresponding name of each feature is shown in Supplementary Table 1.

To quantify the overall *shared information* between pairs of EEG features, which showed significant group differences, by taking not only variables with the largest effect size into account but all variables of the features we used partial least squares correlation (PLSC). For the patients, 55.92% of the pairwise inertias were significant (without correcting for multiple comparisons) and for controls, 40.28%. In general, relative inertias were not very high in both patients (0.254, 0.329, 0.409, for the 25^th^, 50^th^, and 75^th^ percentiles, respectively) and controls (0.305, 0.387, 0.472, for the 25^th^, 50^th^, and 75^th^ percentiles, respectively; **Figure 3**). As in the Pearson’s correlation results, features that showed strong associations were mainly similar features, such as the same network statistics for different connectivity measures in the theta band, for example, at the electrode level: clustering coefficient connectivity estimated with phase locking value (*clust coeff e-plv theta*) and with imaginary part of coherence (*clust coeff e-icoh theta*; ℑ_*relative*_=0.804 and ℑ_*relative*_=0.826, in patients and controls, respectively).

**Figure 3.**
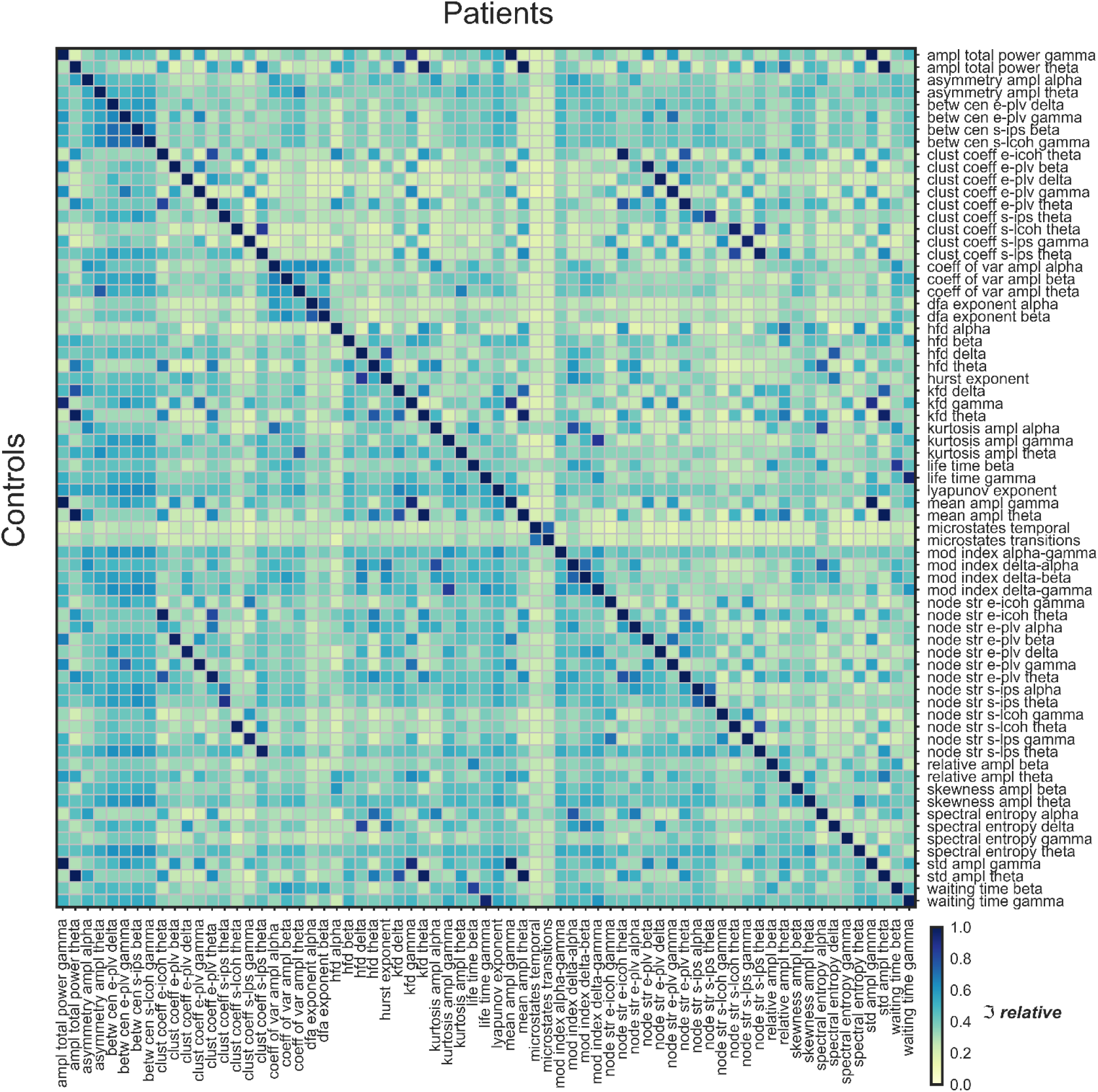
Shared information between the 69 EEG features that showed significant group differences, as measured by the relative inertia (ℑ_*relative*_) computed with partial least squares correlations (PLSC). The relative inertia ranges from 0 (two features are completely unrelated) to 1 (the two features’ values move together by the exact same percentage). Patients’ relative inertias are presented in the upper triangle and controls’ relatives are shown in the lower triangle. A list with the abbreviations and corresponding name of each feature is shown in Supplementary Table 1.

### 3.3. Prediction of psychopathology scores

We evaluated whether EEG features were adequate predictors of psychopathology scores determined by the Scale for the Assessment of Positive Symptoms (SAPS) and the Scale for the Assessment of Negative Symptoms (SANS), which target positive (hallucinations, delusions, bizarre behavior, and positive formal thought disorder) and negative (affective flattening, alogia, apathy, anhedonia, and attention) symptoms, respectively. All 194 EEG features exhibited very weak out-of-sample predictive ability to both the SANS and SAPS scores. Results were very similar for both the linear (i.e., elastic net) and nonlinear (i.e., random forest) models. See Supplementary Table 2 and Supplementary Table 3 for details.

## 4. Discussion

Traditionally, schizophrenia research focuses on a single experimental paradigm and analysis method showing significant differences between patients and controls, and then tries to derive the underlying genetic or neurophysiological causes of the disorder. This approach has been quite successful in the formulation of hypotheses, such as the dopamine hypothesis (Howes and Kapur, 2009), the social brain hypothesis (Burns, 2006), the glutamate hypothesis (Hu et al., 2015), or the dysconnection hypothesis (Friston et al., 2016), just to name a few. Here, we examined to what extent abnormalities quantified by different EEG features correlate with each other. Many of the investigated features were previously linked to different abnormalities of brain processes in schizophrenia, and, here, we reproduced many of these results, such as imbalance in microstates dynamics (da Cruz et al., 2020a; Rieger et al., 2016), decreased long-range temporal correlations in the alpha and beta bands (Nikulin et al., 2012), decreased life-and waiting-times in the beta band (Sun et al., 2014), increased spectral amplitude in the theta band (Boutros et al., 2008), increased connectivity in the theta band at the source level (Andreou et al., 2015; Di Lorenzo et al., 2015), decreased Lyapunov exponent (Kim et al., 2000), among others. With our systematic analysis, we also found abnormalities in EEG features, which, to the best of our knowledge, have not been reported yet, namely, delta-phase gamma-amplitude coupling, range EEG coefficient of variation and asymmetry in the theta and alpha bands, etc. In some way, deeper analysis of each feature may have warranted an in-depth study and a potential publication. However, we did not want to elaborate on these methods individually because we wanted to understand how all EEG features relate to each other in their entirety. The surprising insight from our analysis is that, even though we are probing the same signals from the same participants, we found only weak correlations between the 69 significant features. The only strong correlations were between features that are similar from the outset, thereby resembling test-retests. This suggests that, even though EEG features reveal clear-cut and reproducible atypical patterns in patients, none of the features is truly representative for the disease, but rather that all these features pick up more or less independent aspects of schizophrenia. Hence, the traditional approach of focusing on a single experimental paradigm and analysis method has its limitations. These results remind us that schizophrenia is indeed a very heterogeneous disease, a well-known fact, which is however not always taken seriously enough because, as mentioned above, most research tries to find the one or a few causes of schizophrenia within one well described paradigm by digging as deep as possible into the underlying neurophysiological and genetic mechanisms. In analogy to botany, one may call these approaches “deep rooting” approaches.

We propose that it may be useful to complement these deep rooting approaches with “shallow rooting” approaches, representing schizophrenia within a high-dimensional space, where many tests and analysis outcomes are the basis variables. The outcomes should ideally have large effect sizes, low mutual correlations, and a “flat” factor structure. Whether this is possible is an open question and depends very much on the underlying causes of schizophrenia. On the lowest complexity level, there may be only a few independent causes (or even only one), which were not found yet. Given the heterogeneity of the disease, including abnormalities in the cognitive (Andreasen et al., 1998) but also the skin functioning domain (Messamore, 2012), the causes need to be on a rather general level, likely subcellular, present in all human functioning. Alternatively, schizophrenia may be an approximatively “additive” disease, where many small abnormalities add up to severe symptoms. In an even more complex scenario, only certain combinations of redundant functions, each coming with at least two variants, cause the disease. For instance, if one function is up-regulated and another one down-regulated in an individual, there are no abnormalities. Deficits manifest only when all or most functions are either up- or down-regulated. In such a combinatorial scenario, it would be difficult to find the underlying causes since each variant itself does not lead to a deficit; only certain combinations do. Our correlation analysis provides evidence supporting the more complex/multifactorial scenario, where each feature makes up its own factor and manifests differently in different patients. Since our data shows that each EEG feature is sensitive to roughly independent aspects of schizophrenia, each brain process captured by the analysis methods might be neither necessary nor sufficient to explain important aspects of the disease. Indeed, we found that EEG features showed very weak predictive power to key symptoms of schizophrenia, suggesting that there is little information about individual differences in psychopathology. Our results are an invitation to rethink the current approach in schizophrenia research and suggest that new study designs conflating multiple features from the same and different paradigms might be more adequate. In the next steps, it will be important to find the right set of features, which may stem from EEG recordings but also potentially immunological markers, of which each may contribute with a variety of features. Previous research has shown that combining features improved classification and predictive performance in psychosis studies (Mothi et al., 2019; Yang et al., 2010).

Our study has several limitations. There are demographic differences between patients and controls, which might affect our group comparisons. However, we attempted to minimize these demographic effects by using education as a covariate and gender as factor in the analyses. Similarly, we cannot exclude effects of medication in our results. Nonetheless, we find similar patterns of correlations between EEG features, i.e., weak associations, in both patients and controls, suggesting that if there is an effect of medication, it is small. Further, our sample size is relatively small for achieving reliable estimates of predictive power (Poldrack et al., 2020; Schnack and Kahn, 2016; Varoquaux, 2018).

Our results may explain a deep mystery in schizophrenia research. Schizophrenia has an estimated heritability of 70 to 85% (Burmeister et al., 2008). For example, the chance to also suffer from schizophrenia for monozygotic twins is about 33% when the partner twin has the disease (Hilker et al., 2018). Furthermore, about 0.25 to 0.75% people of a population suffer from schizophrenia and related psychotic disorders (Kessler et al., 2005; Moreno-Küstner et al., 2018; Saha et al., 2005). These values are rather stable across cultures (Simeone et al., 2015). Given that schizophrenia patients have less offspring (Avila et al., 2001; Bassett et al., 1996; Keller and Miller, 2006; MacCabe et al., 2009), this provokes the question why schizophrenia has not been extinguished during the course of evolution (Keller and Miller, 2006; Liu et al., 2019). In the above-mentioned combinatorial scenario with many redundant functions this may simply happen because evolution operates on the individual single-nucleotide polymorphism (SNP) level and not on the combinatorial one. As long as most of the population shows average functioning, there will be no change of the allele distributions. In the additive scenario, evolution may extinct harmful alleles, of which each constitutes only a little risk, very slowly and these may be replaced by harmful de novo mutations (Keller and Miller, 2006). To what extent such considerations hold true will be shown by shallow rooting approaches using a plethora of paradigms and a multiverse of analysis methods. In a nutshell, deep rooting will help to understand the different aspects of the disorder, while shallow rooting will help to better diagnose schizophrenia by finding subpopulations, leading to more personalized treatment.

## Data Availability

The data that support the findings of this study are available upon reasonable request.

## Acknowledgements

We would like to thank Marc Repnow for his comments and Ben Lönnqvist for proofreading the manuscript. M.H.H., E.C., A.B., and M.R. designed the research; M.R. and E.C. performed the research; J.R.C., D.G., W.H.L., and O.F. analyzed data; J.R.C., D.G., O.F. A.B., P.F., and M.H.H. wrote the paper. The code that support the findings of this study are available upon request. This work was partially funded by the Fundação para a Ciência e a Tecnologia under grant FCT PD/BD/105785/2014 and the National Centre of Competence in Research (NCCR) Synapsy financed by the Swiss National Science Foundation under grant 51NF40-185897. The authors declare no competing interests.

